# Regulatory landscape of Alzheimer’s disease variants in human microglia

**DOI:** 10.1101/2025.10.13.25337902

**Authors:** Chia-Yi Lee, Ashvin Ravi, Tzu-Chieh Huang, Briana Wyman, Ziyu Liu, Ariana Chriss, Jack Humphrey, David A. Knowles, Chris Cotsapas, Laura M. Huckins, Towfique Raj, Kristen J. Brennand

**Affiliations:** Departments of Genetics, Yale University School of Medicine, New Haven, CT 06511; Department of Genetics and Genomics, Icahn School of Medicine at Mount Sinai, New York, NY 10029; Department of Psychiatry, Division of Molecular Psychiatry, Yale University School of Medicine, New Haven, CT 06511; New York Genome Center, New York, NY, 10013; Departments of Computer Science, Systems Biology, and Data Science Institute, Columbia University, New York, NY, USA, 10027; Departments of Neurology, Yale University School of Medicine, New Haven, CT 06511; Nash Family Department of Neuroscience, Friedman Brain Institute, Icahn School of Medicine at Mount Sinai, New York, NY 10029

## Abstract

Genome-wide association studies (GWAS) for Alzheimer’s disease (AD) predominantly implicate non-coding genetic variants that are presumed to regulate gene expression. Yet, around half of GWAS variants do not coincide with known regulatory variants, such as expression quantitative trait loci (eQTLs). AD GWAS loci are enriched for microglia-specific regulatory regions, but the causal variants and mechanisms remain unresolved. To connect risk variants to microglial function, we evaluated the regulatory activity of AD GWAS variants, microglial eQTLs, and chromatin accessibility QTLs (caQTLs) by screening 11,550 candidate regulatory sequences (CRSs) with a massively parallel reporter assay (MPRA). Whether prioritized by GWAS or eQTLs, variants are equally likely to predict active CRS and expression-modulating variants (emVars) by MPRA, yet there were clear systematic differences, particularly in that predicted target genes of GWAS loci, but not eQTLs, are enriched for AD relevant biology. Notably, only 7% of all emVars were shared between two distinct sources of human microglia cells, human induced pluripotent stem cell (hiPSC)-derived microglia (iMGLs) and immortalized HMC3 cells, underscoring the importance of cellular context in evaluating regulatory activity. Our findings establish the first large-scale microglial-context MPRA, define a microglial regulome underlying AD genetic risk, and provide a functional framework to refine noncoding variant interpretation.

## Introduction

Over 90% of genome-wide association study (GWAS) loci occur in non-coding regions of the genome and are presumed to regulate the expression of one or more cis-linked genes^1^. Despite the discovery of millions of regulatory variants (expression quantitative trait loci, eQTL) identified in postmortem adult tissues^2^, more than half of GWAS loci do not colocalize with eQTL. The reported differences identified between GWAS loci and eQTLs^3^ may simply reflect methodological biases in their detection. As more recent studies query eQTLs across developmental stages^4^, cell types^5^ and states^6^, and include chromatin accessibility QTLs (caQTLs)^4,7^, the incorporation of context-specificity has improved power to resolve regulatory variants and map causal GWAS loci across diseases.

Despite these advances, the causal mechanisms underlying complex disorders like Alzheimer’s disease (AD) remain unclear. AD has a highly heritable (estimates range from 58%-79%^8^) but complex genetic etiology^9^. Recent genome-wide association studies (GWAS) linked 75 independent loci with AD^10^; 38 of these loci were linked to changes in gene expression^11,12^, more when incorporating microglial splicing QTLs^13^ and chromatin accessibility QTLs^14^. A substantial proportion of AD GWAS loci remain unexplained, although they are highly enriched for microglia enhancer regions^14,15^. Microglia are a particularly heterogeneous^16^ and dynamic cell type^17^, highly impacted by age^18^, and relatively low abundance^19^, making them quite challenging to obtain from large numbers of donors and particularly underpowered in QTL analyses^19^. Consequently, statistical approaches alone have failed to comprehensively capture the functional variants and regulatory mechanisms underlying AD risk.

Massively parallel reporter assays (MPRA) can bridge this gap by systematically and simultaneously testing allelic effects, pairing each variant with a reporter gene bearing unique transcribed barcodes, thereby measuring the regulatory activity of thousands of variants in parallel by high-throughput sequencing^20,21^. To date, most MPRA have been conducted in transformed cell lines^22–24^, with uncertain relevance to human brain cell contexts^24^. Moreover, our recent attempt to map brain disorder GWAS variants in human induced pluripotent stem cell (hiPSC)-derived neurons resolved minimal functional activity of AD GWAS loci^25^, reinforcing the need to test regulatory mechanisms driving AD risk specifically in human microglia.

Here, we performed fine-mapping of the most recent AD GWAS^10^ to prioritize functional variants and integrated these with microglial eQTL and caQTL maps, in total nominating 11,550 risk variants. By conducting MPRA in immortalized human microglia (HMC3) and human induced pluripotent stem cell (hiPSC)-derived microglia-like cells (iMGLs), we conducted the first systematic effort to functionally interrogate Alzheimer’s– and microglia-associated noncoding variants. We tested whether specific variant prioritization approaches (e.g., GWAS, eQTL, caQTL) more effectively predicted MPRA activity and if distinct cellular models (HMC3 and iMGL) better resolved regulatory activity. Our variant-level, gene-level, and network-level analyses deciphered the regulatory outcome of these functional variants and linked genetic risk to cell-type-specific regulatory mechanisms in AD. Altogether, we demonstrated that integrating statistical genetics with high-throughput experimental validation clarified GWAS ambiguity and provided a framework for functional studies in aging-related neurodegeneration.

## Results

### Fine-mapping prioritized functional variants enriched in microglia enhancers

Statistical fine-mapping refined AD GWAS association signals^10,26–28^ and prioritized functional variants (computational schematic, **Fig. 1**). The robustness of four widely used Bayesian and functionally informed approaches (SuSiE^29,30^, FINEMAP^31^, PolyFun^32^ + SuSiE, and PolyFun + FINEMAP) were evaluated. We also assessed whether the choice of LD reference panel influenced results by fine-mapping using pre-computed LD matrices from the UK Biobank as well as those calculated directly from the AD GWAS data^10^. Results were highly concordant across fine-mapping methods and LD panels: 53 of 75 AD risk loci harbored at least one fine-mapped variant (posterior inclusion probability (PIP) > 0.1) across all four methods (95% credible sets) (**Supplementary Table 1**). The distribution of credible sets revealed important distinctions in the strength of fine-mapping resolution: loci with larger credible sets were frequently composed of variants with low PIP that were less likely to contain functional variants, whereas loci with smaller credible sets were enriched for high-PIP variants with stronger support for specific functional candidates (**Fig. 2A**). A modest fraction (16.5%) of variants were consistently prioritized across all fine-mapping methods (**Fig. 2B**). Our analysis resolved well-characterized (e.g. rs75932628)^33,34^ and novel variants at the *TREM2* locus, whereas rs6733839 emerged as the sole credible functional variant at the *BIN1* locus^35–37^ (**Fig. 2C**). Fine-mapped variants (the 95% credible set) were disproportionately located in microglial enhancers (**Fig. 2D-E; Supplementary Fig. 1A, Supplementary Table 2**) and other immune cell types^38^ (**Fig. 2F; Supplementary Fig. 1B**), being enriched for genes linked to immune regulation and microglial biology (e.g., *PLCG2*, *SPI1*, and *RASGEF1C*), and showing stronger enrichment for microglial enhancers than lead AD GWAS variants (**Fig. 2G; Supplementary Fig. 1C**). Altogether, fine-mapping substantially improved localization of AD risk variants, strengthening functional interpretation within microglia.

**Figure 1.**
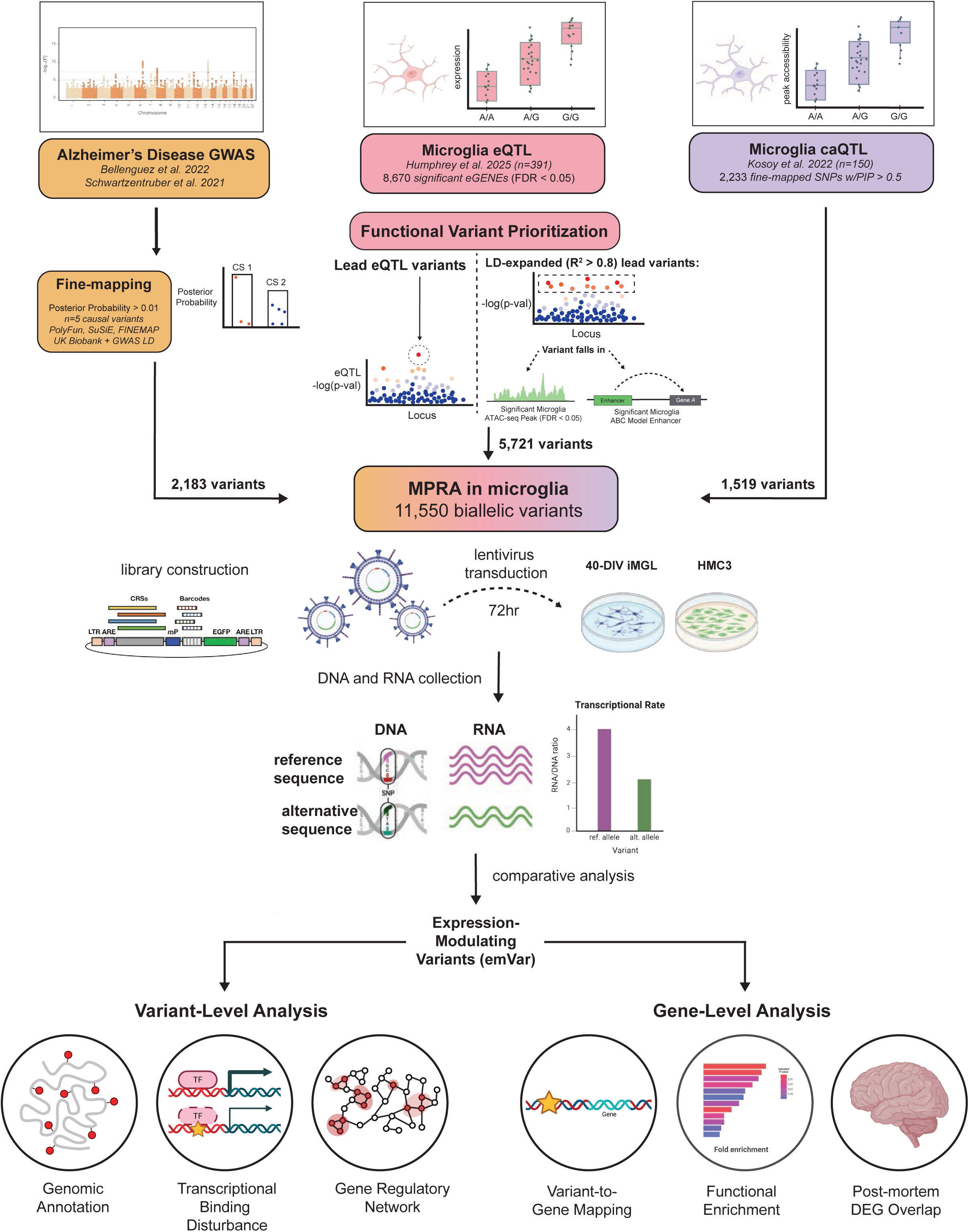
Overview of MPRA library design and experimental workflow. Fine-mapped Alzheimer’s disease (AD) GWAS variants, microglial eQTL, and caQTL were included in the MPRA library. Each construct contained a candidate regulatory sequence (CRS), a unique barcode, and a reporter gene. The library was packaged into lentivirus and introduced into microglial cell models (HMC3 and iPSC-derived microglia). DNA and RNA were extracted from the same cultures to quantify allele-specific transcriptional activity by comparing RNA/DNA ratios between reference and alternative alleles. Expression-modulating variants (emVars) were subsequently analyzed at both the variant and gene levels.

**Figure 2.**
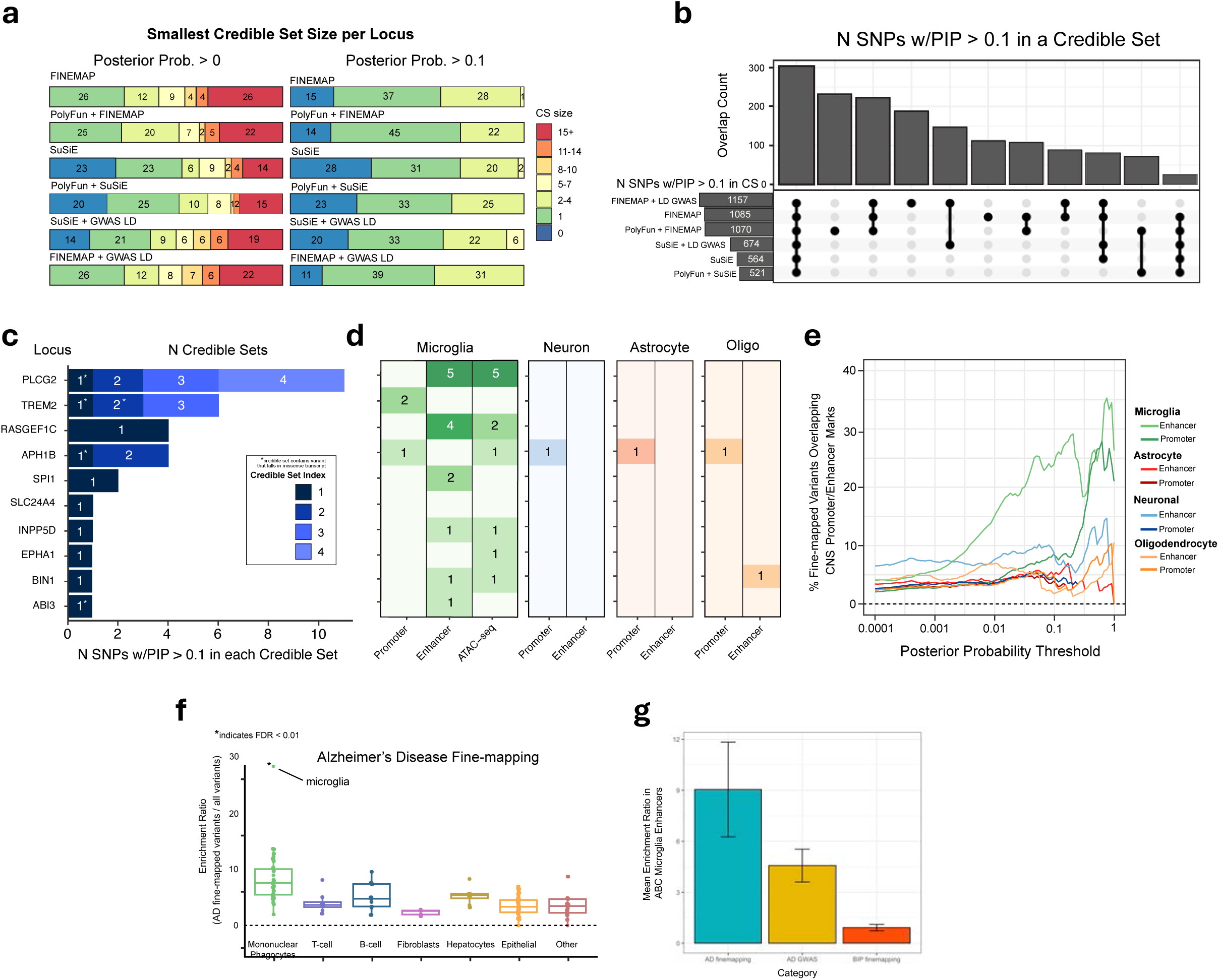
Summary of Fine-mapped Variants in Alzheimer’s Disease (AD). (a) Size of the smallest credible set per locus for all variants and variants with posterior probability > 0.1 (b) Upset plot showing the number of fine-mapped variants overlapped across methods (c) Fine-mapped results across 10 subset AD GWAS loci (d) Number of fine-mapped SNPs within each locus that overlap promoter and enhancer marks in microglia, neurons, astrocytes, and oligodendrocytes (e) Fraction of fine-mapped variants above a given PP threshold that overlap promoter or enhancer marks across cell types in the central nervous system (f) Enrichment ratio of AD fine-mapped variants within enhancers identified by the Activity-by-Contact (ABC) model from 132 cell types (g) Enrichment ratio of AD fine-mapped variants, lead AD GWAS SNPs, and fine-mapped variants in bipolar disorder (BD) within ABC microglia enhancers

### hiPSC-derived microglia, more so than HMC3s, capture AD-relevant regulatory programs

A comprehensive set of candidate AD-associated functional variants was prioritized for MPRA assessment (**Fig. 1**). First, all fine-mapped variants from AD GWAS^10,28^ studies, together with the variants identified in the most recent AD fine-mapping study^10,26,27^ (PIP > 0.01; total variants = 2,183) were included. Second, lead microglia eQTL variants (n = 5,721) from a meta-analysis of 391 donors^13^ were incorporated, along with variants in strong LD (R^2^ > 0.8) with these lead eQTLs. We then filtered this set to variants overlapping either significant primary microglia ATAC-seq peaks (n = 1,519) or microglia Activity-By-Contact (ABC) enhancer–gene links (n = 730). Third, microglia caQTL variants with high confidence fine-mapping (PIP > 0.5; n = 2,074) were included^14^. Together, this integrative approach yielded a consensus set of 11,550 putative functional variants, representing high-confidence regulatory candidates for AD (**Supplementary Table 3**). To test both alleles of each variant, we synthesized 23,100 candidate regulatory sequences (CRSs), along with 844 scrambled and positive/negative controls (**Fig. 1**).

In our MPRA library, each CRS was linked to a unique barcode (barcodes per CRS: mean = 119, median = 106); transcriptional activity was quantified as the abundance of barcode RNA relative to its corresponding DNA [log₂(RNA/DNA)]. The MPRA library was transduced into mature iMGLs (40 DIV; two control donors, three replicates from a male and four from a female donor, 72 hours) or into HMC3 cells (three biological replicates each, 72 hours). MPRA coverage was high, with approximately 95% of CRSs detected in both microglial models, and strong inter-replicate correlations among DNA counts, RNA counts, RNA/DNA ratios (r = 0.60-0.88), and barcode counts (**Supplementary Fig. 2**). To ensure reliable activity estimates, CRSs with <10 barcodes or with low RNA/DNA ratio were excluded (**Fig. 3A**). Expression-modulating variants (emVar) were defined as sites with significant allele-specific activity (p_FDR_ < 0.05) in iMGLs (**Supplementary Table 4**) and HMC3 (**Supplementary Table 5**).

**Figure 3.**
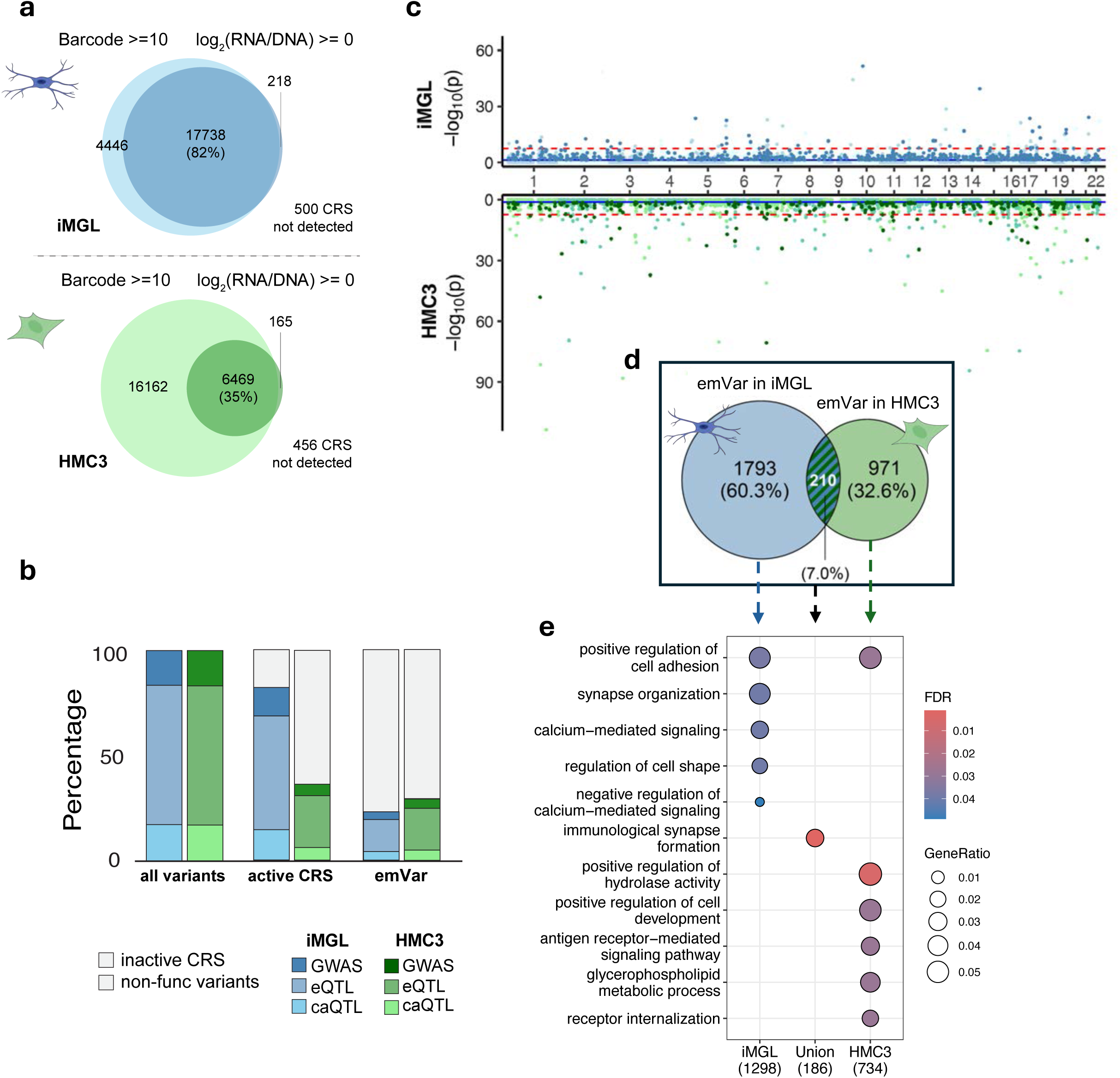
Clear distinction in CRS and frVar across Prioritization Methods and Cell Types. (a) Total number of Candidate Regulatory Sequences (CRS) in the library that passed filtering for low barcode counts and low CRS activity. (b) Percentage of CRS and Expression-Modulating Variants (emVar) categorized by their method of prioritization. “All variants” refers to all variants in the tested MPRA library, and “active CRS” refers to variants with detectable transcriptional activity. (c) Genome-wide distribution of emVar in iPSC-derived microglia-like cells (iMGL) and HMC3 cells. The blue line indicates FDR 5 × 10^-2^, and the red dotted line indicates FDR 5 × 10^-8^. (d) Overlap of emVar identified in iMGL and HMC3 cells. (e) GO-term enrichment analysis on the intersect and union of emVars-lnked nearest genes from two cell-lines.

We first assessed whether genetic evidence predicted MPRA activity. Overall, neither method of variant prioritization, GWAS or QTL-based, better predicted CRS activity or the proportion of emVars by MPRA; the fraction of active CRSs did not differ among GWAS, eQTL, or caQTL variants (**Fig. 3B**). Likewise, across finer-grained categories nominated as lead GWAS variants, fine-mapped GWAS credible sets, lead eQTLs or their LD proxies, or caQTLs methods, the likelihood that a variant exhibited transcriptional activity or allelic regulatory effects was comparable (**Supplementary Fig. 3A**). However, multiple annotations (i.e., in the ∼6% of variants with two or more of GWAS, eQTL, and caQTL; **Supplementary Fig. 3B**) better predicted emVar status (β = 0.18, p = 0.03; **Supplementary Fig. 3C**), suggesting that these datasets capture largely distinct but partially convergent subsets of functional variants.

To contextualize cell-type specific regulatory differences, we compared the transcriptomic states of the two microglial models. Transcriptomic analyses revealed substantial differences between the cell types, highlighted by principal component analysis (PCA) (**Supplementary Fig. 4A**). iMGLs more closely resembled primary microglia, whereas HMC3 expression revealed increased cell-cycle (chromosome segregation, nuclear division, and mitosis) and decreased immune and metabolic pathways (immune cell activation and immune response) (**Supplementary Fig. 4B,C**).

These transcriptional distinctions were mirrored in the MPRA results. In total, 2,003 emVars were identified in iMGL and 1,181 in HMC3 (**Fig. 3C**), with minimal overlap between cell types: only 21.5% of HMC3 signal replicated in iMGL (**Fig. 3D**). Gene Ontology (GO) enrichment of the predicted emVar target genes revealed that iMGLs were more enriched for hallmarks of microglial function and neuron-microglia communication (e.g., synapse organization, regulation of cell shape and adhesion, and calcium-signaling pathways) (**Fig. 3E, left**). In contrast, HMC3 enrichments emphasized generic signaling and metabolic programs (**Fig. 3E, right**). Although the overlapping signals centered around immunological synapse formation (**Fig. 3E, middle**), the largely divergent regulatory programs between iMGL and HMC3 underscore the importance of assaying variant function in a physiologically relevant microglial model.

As iMGLs showed greater transcriptomic similarities to primary microglia, exhibited the strongest regulatory signals, and had a greater number of emVar than HMC3, we focused downstream analyses on iMGL.

### iMGL emVars concentrate in promoter-proximal features and regulate disease-relevant TF

Fine-mapped GWAS SNPs (high posterior inclusion probability (PIP) ≥ 0.5) were indeed more likely to modulate allele-specific transcriptional activity compared to low-PIP variants (0.2 > PIP ≥ 0.1) (pairwise Wilcoxon test, BH-adjusted p = 0.018) (**Fig. 4A**). iMGL emVar were significantly enriched in canonical regulatory elements (**Fig. 4B**), and MPRA-positive emVars (p_FDR_ < 0.05, relative to MPRA-negative, p_FDR_ > 0.05) were enriched in microglial promoter regions (OR = 1.28, p = 0.02) (**Fig. 4C**) and iMGL ATAC-seq peaks (**Supplementary Fig. 5**).

**Figure 4.**
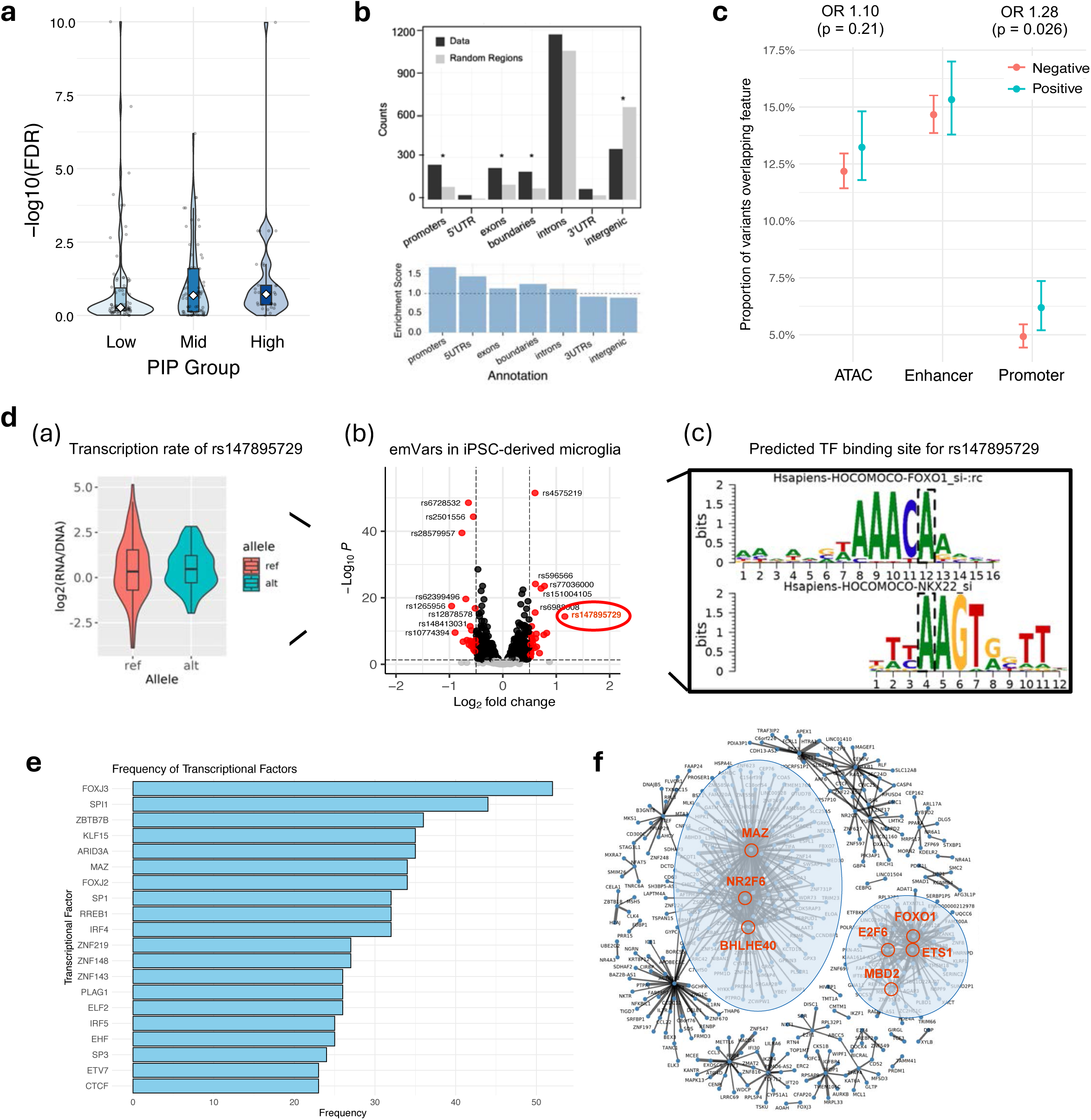
Function of emVar and Their Disease-Relevant Enrichment. (a) Distribution of significance by PIP group (low 0.2 > PIP ≥ 0.1; mid 0.5 > PIP ≥ 0.2; high PIP ≥ 0.5). (b) Enrichment of emVars in ENCODE genomic annotations and fold-change comparison between emVar and all tested variants (allVar). Enrichment reflects observed-to-control frequency, and the enrichment score is the ratio for emVars versus all MPRA variants. (c) Enrichment of emVar in microglial annotation. (d) a. Distribution of transcriptional activity for reference (REF) vs alternate (ALT) alleles at the example variant. b. Volcano plot of emVars activity across tested variants with example variant rs147895729 circled. c. Transcription factor (TF) binding motif disrupted by the example variant. (e) TF binding site prediction for CRSs containing emVar. (f) TF-centered gene-regulatory network.

The fold-change in transcriptional activity between alternative and reference alleles was generally modest, with most variants exhibiting less than two-fold differences (**Fig. 4D**). Mechanistic interpretations of selected confidence emVars were clear: for example, an emVar at rs147895729 revealed allele-specific effects predicted to disrupt transcriptional factor (TF) FOXO1 and NKX2.2 binding (**Fig. 4Dc**). Extending beyond this example, approximately 50% of CRSs overlapped predicted TF binding sites (**Fig. 4E**), consistent with promoter-proximal regulation (**Fig. 4B**). Top enriched TFs revealed links to neurodegeneration^39–41^ and neurodevelopment^42–46^; for example, SPI1, a key myeloid regulator^47^ linked to regulation of AD risk variants and amyloid-β pathology^48,49^. A microglial gene regulatory network (random-forest approach^50^) constructed from co-expression between TF and target gene expression profiles (Fig. 4F) resolved two programs: immune/inflammatory (MAZ^51^-NR2F6^52^-BHLHE40^53^) and DNA-damage/stress-response program (FOXO1^54^-E2F6^55^-ETS1^56^-MBD2^57^), the latter already implicated in AD^58–60^, reinforcing the relevance of stress-response pathways to microglial dysfunction in AD.

### MPRA reveal complementary biology across prioritization strategies

We applied three variant-to-gene mapping approaches: microglial Activity-By-Contact (ABC)^61^, microglial eQTLs, and nearest-gene mapping (**Fig. 5A-B**). Overall, gene mapping of emVars captured homeostatic, inflammatory, and metabolic processes rather than converging on a single pathway (**Fig. 5C**). When stratifying variants by prioritization method, GWAS fine-mapped emVar target genes aligned with Alzheimer’s disease mechanisms and were enriched for amyloid biology and immune/inflammatory responses (**Fig. 5C, left**). In contrast, active caQTL targets were enriched for neurodevelopmental processes (**Fig. 5C, right**), whereas active eQTL targets showed no significant functional enrichment (**Fig. 5C, middle**), consistent with previous reports^3,4^. Together, these results underscore the phenotypic complexity of GWAS loci, with complex cell-type-specific and context-dependent regulatory landscapes.

**Figure 5.**
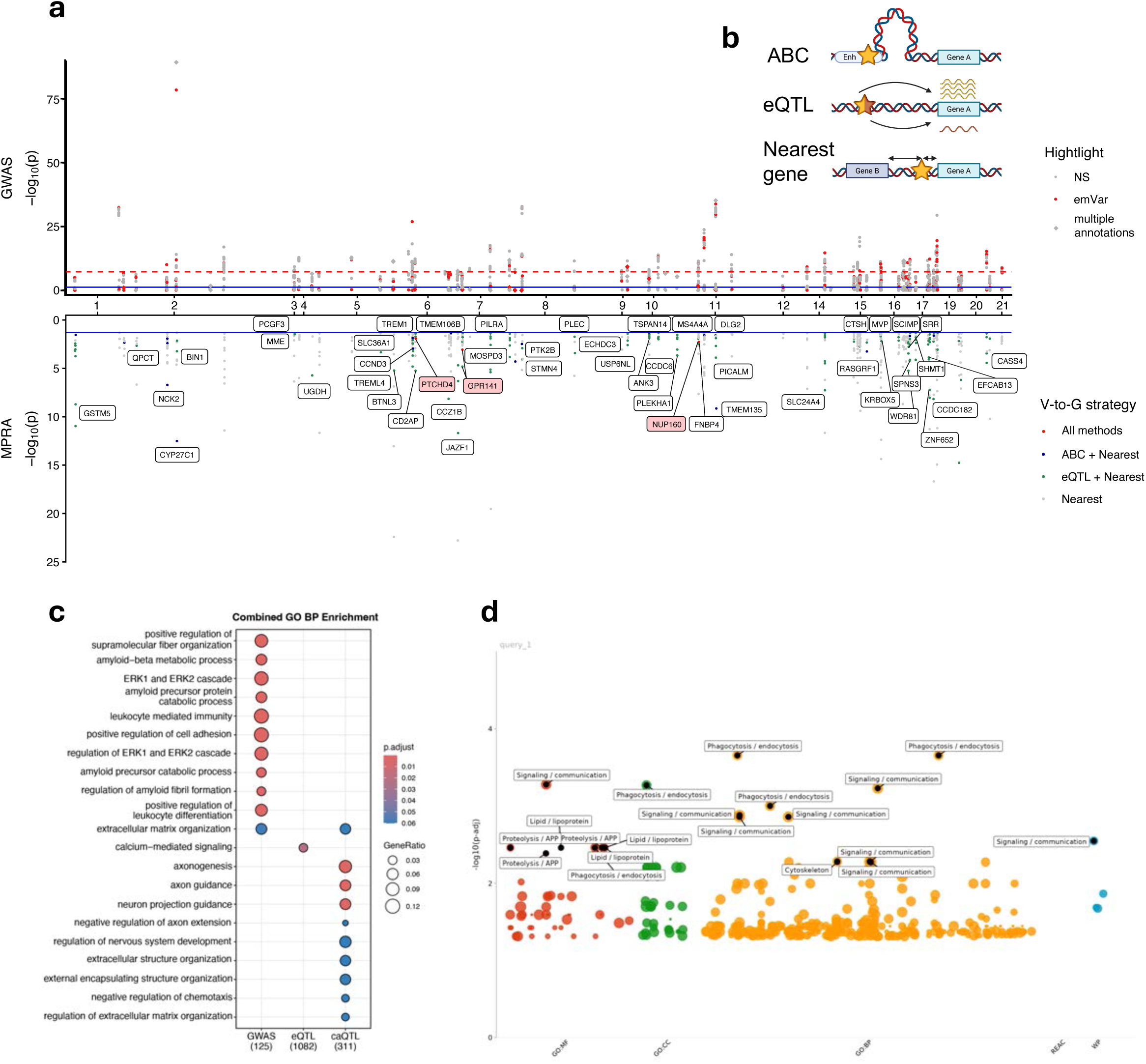
Variants prioritized by different methods reveal distinct biological regulation. (a) Alzheimer’s disease GWAS loci: the upper Manhattan plot showing GWAS results from Bellenguez et al.; red points indicate variants with significant regulatory activity identified by MPRA, and purple points highlight variants supported by multiple variant prioritization method. The lower Manhattan plot shows iMGL MPRA results; points are colored based on variant-to-gene (V-to-G) mapping strategies, and genes supported by multiple strategies are labeled. (b) Schematic overview of the variant-to-gene mapping framework. (c) Gene Ontology (GO) term enrichment analysis of nearest genes from each variant prioritization methods. (d) Pathway enrichment of overlapping genes between emVar-linked and AD differentially expressed genes.

In total, we identified 50 high confidence emVar-linked AD genes mapped by two or more variant-to-gene methods (Fig. 5A). In addition, for each AD GWAS locus, we comprehensively annotated and compared the predicted gene targets of all prioritized variants relative to functionally active emVar variants (**Supplementary Fig. 6-7**); emVar-linked genes were preferentially expressed in a subpopulation of post-mortem brain microglia associated with late-stage AD^62^ (**Supplementary Fig. 8A**) and significantly concordant with microglia (but not whole brain) eQTLs, emphasizing the importance of considering cell-type and state in interpreting GWAS variant regulatory activity (**Supplementary Fig. 9**). The target genes of AD GWAS-prioritized emVars overlapped with differentially expressed genes (DEGs) identified in post-mortem AD microglia (permuted p= 0.0015; **Supplementary Fig. 8B**), enriched for phagocytosis-related processes (endocytosis, vesicle-mediated transport) and cellular communication pathways (signal transduction, cell communication, signaling) (**Fig. 5D**).

### Evaluation of Regulatory Variant Candidates at the *TSPOAP1* Locus

The power of MPRA lies in its ability to directly identify functional regulatory variants, thereby overcoming the limitations of purely *in silico* predictions. For example, the *TREM2* R47H missense variant (rs75932628), described earlier in the fine-mapping section, was further validated experimentally as an emVar in iMGL. Beyond such well-characterized loci, we also identified emVar in previously understudied regions. *TSPOAP1* represents a novel and largely uncharacterized locus associated with AD. Statistical fine-mapping identified six putative causal variants within the credible set, with GWAS SNP rs2680700 showing the highest PIP (**Fig. 6a**). Yet, a second credible set contained many variants with lower PIPs (**Fig. 6b**); this region overlapped multiple microglia-relevant annotations such as promoters, enhancers and ATAC-seq peaks, highlighting its potential regulatory importance (**Fig. 6c**). Colocalization analysis suggested a potential shared causal signal between the AD GWAS and microglial eQTL (PP.H4 = 0.55), although the evidence did not strongly exclude alternative models of distinct signals (PP.H3 = 0.15) (**Supplementary Fig. 10**); indeed, MPRA identified two emVars (rs2526377: PIP=0.163 and rs116939255: PIP=9.44e-08) at *TSPOAP1* locus (**Fig. 6d**), neither of which is the highest-PIP variant. This suggests that the GWAS and eQTL associations at this locus may arise from distinct regulatory mechanisms.

**Figure 6.**
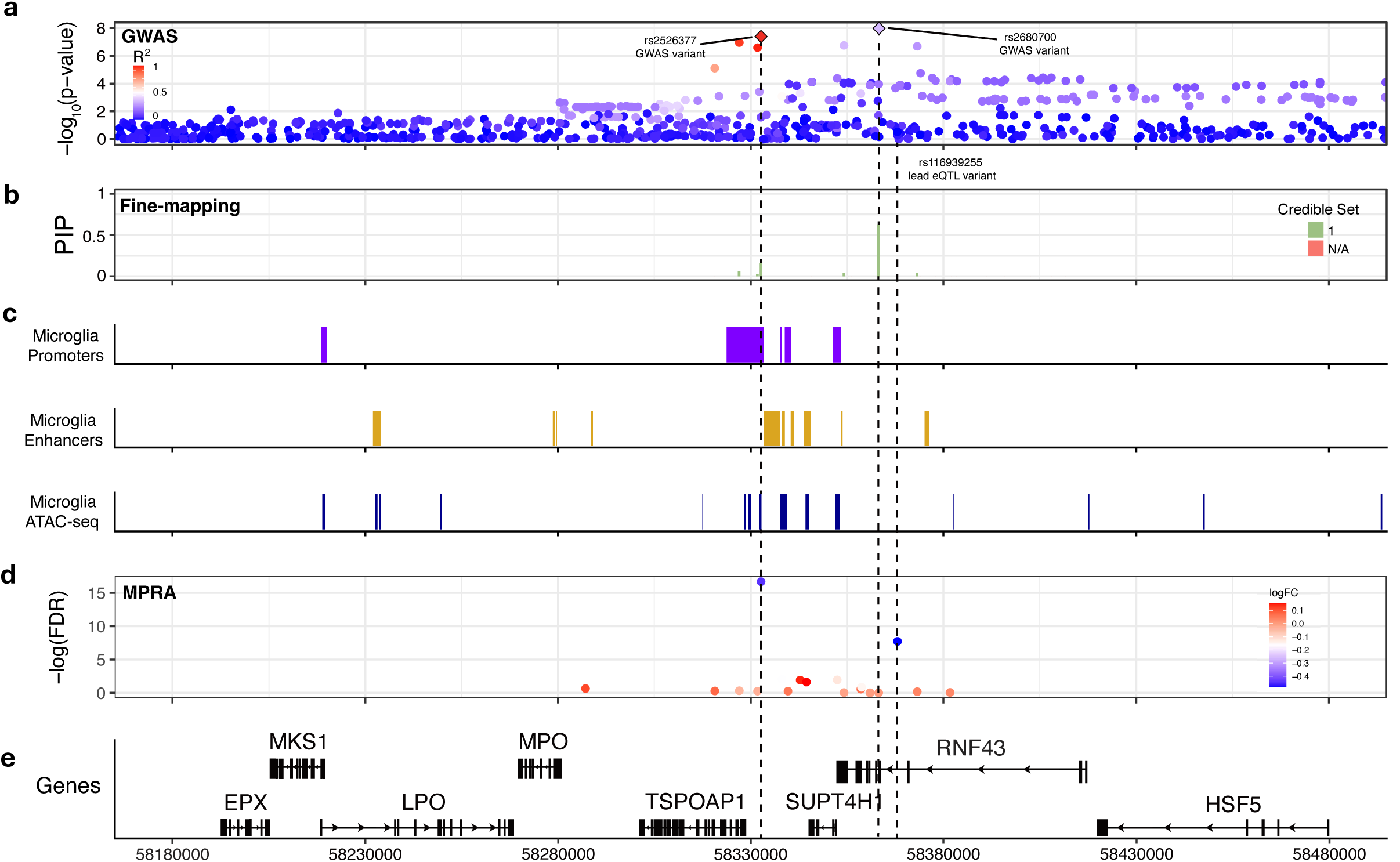
Fine-mapping and MPRA results for AD GWAS *TSPOAP1* Locus. This composite plot integrates multiple layers of genetic and functional evidence for one representative AD locus: (a) GWAS summary statistics from Bellenguez et al., colored by linkage disequilibrium with the lead SNP; (b) SuSiE fine-mapping results showing two credible sets; (c) chromatin accessibility and regulatory annotations, including primary microglia promoter and enhancer peaks and iMGL ATAC-seq peaks; (d) MPRA activity results for variants within the locus; and (e) gene models for all genes located in this region.

## Discussion

We established a scalable experimental framework to link genetic risk variants to regulatory function, reporting the first systematic screen to examine the functional impact of AD-associated variants and QTL variants in human microglia. We assembled a list of candidate functional variants prioritized by statistical fine-mapping, eQTL, and caQTL studies in human microglia, and tested these variants for regulatory activity in immortalized HMC3 and mature iMGL. By applying MPRA in both immortalized HMC3 and mature iMGL, we comprehensively characterized the regulatory potential of variants identified from GWAS, eQTL, and caQTL studies across two cellular contexts. emVar were significantly enriched in microglial regulatory regions, overlapped with TF binding sites, and revealed two core regulatory modules. Overall, GWAS-prioritized variants were enriched for amyloid-related pathways, caQTL-prioritized variants emphasized neural development, and eQTLs lacked substantial enrichments. We demonstrate that MPRA pinpoint putative functional variants in the novel AD loci and resolve multiple overlapping signals in a cell-type-specific manner. Altogether, our results inform the design of subsequent MPRA, reinforce distinct biased between GWAS and QTL studies, and highlight the necessity of measuring activity in the most physiologically relevant cell type available.

HMC3 is a widely used cellular model for human microglia^22–24^, valued for its primary microglial origin and ease of maintenance. Yet, HMC3 cells are less reactive than typical immune cells^24^ and show limited evidence of hallmark microglial functions^24,63^. Consistent with these limitations, only 21.5% of emVars identified in HMC3 replicated in iMGL, reflecting substantial cell-type-specific regulatory differences. We propose that iMGL provides a more informative cellular context for dissecting regulatory mechanisms in microglia. Consistent with the previous report that most early developmental eQTLs were also present in adult tissue^4^, despite their relatively early developmental stage, MPRA results from hiPSC-derived iMGLs showed strong concordance with eQTLs identified in adult tissues.

A key question in statistical genetics is whether integrating multiple variant prioritization methods better predicts functional activity^64–67^. Being composed of ∼25% AD GWAS-, ∼50% eQTL-, and ∼25% caQTL-prioritized variants, our MPRA revealed no clear advantage to any single prioritization strategy. Yet, the limited overlap (∼6%) between approaches reinforces recent findings suggesting that GWAS-, eQTL-, and caQTL-prioritized regulatory variants capture distinct yet related biological processes^3,4^. Of note, GWAS loci often overlap distal regulatory elements with small effects, which are particularly challenging to identify using eQTL assays^4^. Likewise, MPRAs evaluate variants outside their endogenous context, only testing ∼200 base pairs of sequence at a time, omitting larger regulatory regions, and not testing distal regulatory activity. These technical limitations might underly our observed enrichments in transcription factor binding sites, given their short-range sequence-based regulation.

Our results offer a new perspective on the genetic influence on AD and microglia biology. As a complex trait, AD risk seems to be distributed across a broad range of regulatory contexts^4^; particularly given that distinct microglia states and spatial–temporal heterogeneity are linked to AD pathology^9^, testing variant activity across dynamic states will be necessary. Improved mapping of GWAS loci to causal variants will inform our understanding of disease mechanisms.

## Methods

### GWAS Data

We used the European Alzheimer’s Disease Biobank (EADB) Stage I summary statistics for functional fine-mapping^10^. This GWAS analysis was based on 39,106 clinically diagnosed AD cases, 46,828 cases of proxy-AD and related dementia, and 401,577 controls. 21,101,114 variants passed quality control and variants below p = 1 x 10^-5^ were selected. Conditional analyses (GCTA-COJO) revealed 75 loci and 82 lead variants. 33 of the loci were previously associated with AD and 42 loci were newly reported in this GWAS. We also used fine-mapping results from a meta-analysis^28^ of 75,000 AD cases and 420,000 controls from the most recent GWAS analysis^26^ as well as samples from the UK Biobank. Conditional analyses identified 37 risk loci, where 33 were previously reported and 4 were newly reported in this meta-analysis. Fine-mapping results using PAINTOR^69^ and FINEMAP^31^ methods were provided by this study. To maintain consistency between fine-mapping results using EADB data, we select all fine-mapped variants from this study with a posterior probability greater than 0.01 for further analysis^28^.

### Fine-mapping Methods

For each lead signal/locus, we gathered all SNPs within 2-Mb windows (± 1 Mb flanking the lead GWAS SNP) and filtered out SNPs with a minor allele frequency (MAF) < 0.001. We focused on common variants to maximize the relevance of these results to a larger proportion of the AD population.

LD correlation matrices (in units of r) were acquired for each locus from the UK Biobank (UKB) reference panel, pre-calculated by Weissbrod et al.^32^. In addition, LD correlation matrices were calculated for each locus using the matched sample-level data from the EADB GWAS^10^. Any SNPs that could not be identified within the UKBB + GWAS LD reference were removed from subsequent analyses.

Statistical fine-mapping was performed on each locus separately with FINEMAP^31^ and SuSiE^29^. Functional fine-mapping was performed using PolyFun^70^ + SuSiE and PolyFun + FINEMAP, both of which compute SNP-wise heritability-derived prior probabilities using an L2-regularized extension of stratified-linkage disequilibrium (LD) Score (S-LDSC) regression^71,72^. For PolyFun + SuSiE, we used the default UK Biobank baseline model composed of 187 binarized epigenomic and genic annotations. In all subsequent analyses presented here, SNPs that fall within the HLA region were excluded due to the particularly complex LD structure.

While the specifics of each fine-mapping model differ from tool-to-tool, they are united by several key features: 1) they are Bayesian models; 2) they provide the posterior inclusion probability (PP) that each SNP is causal, on a scale from 0 to 1, and 3) they provide credible sets (CS) of SNPs that have been identified as having a high PP of being causal, which we have set at a threshold of PP ≥ 0.95 for all tools. We only included tools that met the following criteria: 1) can take into account LD and 2) can operate using only summary statistics. For all fine-mapping models, we set the (maximum) number of causal SNPs to five.

### ABC Model Bootstrap Analysis

We used Activity-by-Contact (ABC) model results from 131 different cell types^38^. Significant enhancers were determined by filtering out all enhancer-gene pairs that have an ABC score less than 0.02 in each cell-type (threshold determined by the original authors). We also use ABC model results that used microglia ATAC-seq and Hi-C data from the Kosoy et al.^14^ paper, in order to determine cell-type specific enrichment in microglia compared to the 131 other cell types. A bootstrap test was performed to assess enrichment of AD fine-mapped variants in cell-type specific enhancers identified by ABC. We intersected variants with PP ≥ 0.1 in noncoding credible sets with ABC enhancers. For each cell-type, we calculate an enrichment ratio which is the number of fine-mapped variants that overlap ABC enhancers divided by the same number of randomly sampled common variants from the 1000 Genomes Project. We defined a cell-type as significantly enriched if the adjusted p-value (FDR) was <0.01.

### MPRA Library Construction

The MPRA library preparation was adapted from the published lentiMPRA protocols^73^. Briefly, a library of candidate regulatory sequences and adaptor sequences used for library construction was synthesized as HiFi oligonucleotides library (∼230 bp) from Agilent Technologies. These oligonucleotides underwent two rounds of PCR for amplification and addition of random 15-bp barcode sequences. The PCR products are cloned into a lentiviral vector upstream of a minimal promoter and an eGFP reporter gene with NEBuilder HiFi DNA Assembly Master Mix (NEB E2621X). Each candidate sequence was associated with an average of around 100 unique barcodes to enable accurate quantification of reporter activity. The fragments were inserted into the pLS-SceI vector (Addgene #137725) between SbfI and AgeI enzyme cutting site and then transformed into NEB 10-beta Electrocompetent E. coli cells (NEB C3020K) via electroporation. Bacterial stock containing a mixture of the library were grown overnight on Carbenicillin-positive plates (Teknova L5010) and maxiprepped for plasmid collection (Invitrogen K210017). CRS and barcode association are identified from the plasmid by Noveseq.

### Microglia cellular culture

HMC3 is a cell-line purchased from American Type Culture Collection (ATCC CRL-3304) and cultured in the Minimum Essential Medium media (Gibco 11095080) with 10% FBS. hiPSC-derived microglia were differentiated from two control iPSC lines (#3182-XX and #2607-XY)^74^. The protocol was adapted from Abud^75^ and McQuade^76^ et al. The media from StemCell Technologies were used to make hematopoietic stem and progenitor cells (HSPC) and then differentiated into microglia. Briefly, hiPSC was grown in sparse aggregates with hematopoietic media (STEMdiff™ Hematopoietic Kit #05310) for 12 days. Non-adherent cells CD43+ HSPC were further plated with media (STEMdiff™ Microglia Differentiation Kit #100-0019) supplemented with three key cytokines critical for microglia proliferation and maintenance. The differentiating microglia were sub-cultured every 8 days and were collected on day 24 after plating HPSCs into the microglial differentiation media. The final maturation step is achieved by the addition of neuronal and astrocytic factors into the maturation media (STEMdiff™ Microglia Maturation Kit #100-0020) and the matured iMGLs can be collected after another 4 days of culture. The iMGLs were stained with markers to confirm its identity.

### RNA and ATAC sequencing analysis

RNA-sequencing (RNA-seq) was performed in triplicate to characterize the transcriptional profile, and ATAC-sequencing (ATAC-seq) was used to analyze the chromatin accessibility. Both RNA and ATAC sequencing are done in the day 8 matured iMGLs. Cells for RNAseq were harvested in Triton X-100 (Sigma-Aldrich) and libraries were prepared using KAPA mRNA HyperPrep kit. Paired-end sequencing reads (150bp) were generative with NovaSeq. Raw reads were aligned to hg38 using STAR aligner (v2.7.11a)^77^ and gene-level expression matrices were generated by featureCounts^78^ (Subread v2.0.3^79^) with GRCh38 genome and GENCODE^80^ v40 annotation. Genes with over 10 counts per million (CPM) in all samples were retained and the lowly expressed genes were otherwise removed to ensure stable expression profiles. Cells for ATAC were harvested and freshly frozen in 100% FBS. DNA library was prepared with the Active Motif ATAC-seq kit. Paired-end sequencing reads (150bp) were generative with NovaSeq. Adaptors in raw reads were cut with cutadapt (v5.0)^81^ and quality-checked with FastQC (v0.12.1)^82^. The preprocessed fragments were aligned using Bowtie2 (v2.4.2)^83^. Mitochondria reads were removed and BAM files were created, sorted, and indexed with SAMtools (v1.16)^84^. Peaks were called with MACS2 (v2.2.9.1)^85^ and mapping rates were checked with a custom python script. Peaks were intersected between replicates and overlapping peaks were merged by BEDTools (v2.30.0)^86^.

### MPRA lentivirus Transduction

The lentiviral MPRA library was packaged into second-generation lentiviral particles by co-transfecting HEK293T cells with the library plasmid together with psPAX2 and pMD2.G. Virus production was performed by the Boston Children’s Hospital Viral Core.

For MOI calculation, viral titers were calibrated in parallel using a control lentivirus (Addgene #137724). Following transduction with serially titrated control virus, genomic DNA was isolated and integrated viral copies were quantified by qPCR with background signal subtracted to estimate functional titer and determine the volume of virus required to achieve the desired MOI for MPRA experiments.

For MPRA transduction, iMGLs were transduced on day 5 of maturation, and HMC3 cells were transduced at ∼70% confluency. To achieve an average multiplicity of infection (MOI) of ∼50, 30 µL of virus was used per 1 × 10^6^ iMGLs and 42 µL of virus was used per 1 × 10^6^ HMC3 cells. On average, 5 × 10^6^ cells were used per iMGL replicate and 1 × 10^7^ cells were used per HMC3 replicate. Genomic DNA and mRNA were harvested from the same cultures for barcode sequencing.

### MPRA sequencing data analysis

The plasmid library was sequenced on a MiSeq instrument to link each candidate regulatory sequence (CRS) with its barcode. Pooled genomic DNA and RNA from the same culture after the MPRA experiment were sequenced on a NovaSeq. Sequencing data were processed using MPRAflow (version 2.0)^73^ for initial read alignment, barcode counting, and CRS-barcode association. Only CRSs with at least 10 unique barcodes were retained. For each variant, incomplete reference-alternate pairs were removed. CRSs with a transcriptional rate below log2 RNA/DNA ≤ 0 were excluded, ensuring that only active constructs were analyzed.

We analyzed the resulting DNA and RNA count matrices using MPRAnalyze (version 1.24.0)^87^, which employs a generalized linear model with a negative binomial distribution to jointly model RNA and DNA counts while accounting for library size and replicate depth. We estimated normalization factors and scaled both DNA and RNA libraries per replicate. Scrambled sequences were defined as controls and used to calibrate the null model in the scale mode. For allelic comparisons, we modeled RNA expression under reference vs. alternative allele and tested for allelic effects using a likelihood-ratio test. The resulting log₂ fold-change (logFC) represents the transcriptional difference of the alternate relative to the reference allele. Multiple testing correction was applied using the Benjamini–Hochberg method, and variants were called expression-modulating variants (emVar) if the allelic test reached FDR < 0.05 within each cell model.

Each MPRA was performed in three biological replicates for iMGLs derived from two donors (donor #3182-XX, n = 4) and for the HMC3 cell line (n = 3). Replicate correlations for iMGLs across donors ranged from r = 0.88–0.98 for RNA, r = 0.85–0.96 for DNA, and r = 0.55–0.85 for RNA/DNA ratios. For HMC3, replicate correlations were r = 0.96 for RNA, r = 0.94–0.98 for DNA, and r = 0.80–0.90 for RNA/DNA ratios. Counts were aggregated across an average of ∼100 barcodes per CRS prior to modeling. emVar were defined at the variant level, and the identified emVar were used as input for downstream analyses.

### Variant functional annotation

The general ENCODE genomic annotations — including promoter, UTR, exon, intron, and exon-intron boundary information — were obtained from Bioconductor UCSC genomic features database (TxDb.Hsapiens.UCSC.hg38.knownGene)^88^ and annotated with R package annotatr (v1.32.0)^89^. Both the entire library and functional subset were compared with randomly selected variants from the same chromosome with the randomize_regions function. All results of enrichment are compared using a Chi-squared test and adjusted for multiple testing using Benjamini-Hochberg correction.

Promoter and enhancer annotations for microglia were derived from primary human microglia^35^. Active promoters were identified by H3K27ac and H3K4me3 peaks overlapped; active enhancers were identified by H3K27ac peaks that were outside of H3K4me3 peaks. ATAC-seq peaks were obtained from matured iPSC-derived microglia. Enrichment differences between MPRA-positive and MPRA-negative variants were assessed using a Fisher’s exact test.

### GO-term enrichment analysis

All Gene Ontology (GO) annotations were obtained from org.Hs.eg.db (v3.19.1)^90^, and enrichment analyses were performed using the clusterProfiler R package (v4.12.0)^91^. Nearest-gene mappings of functional variants were used as input, based on Ensembl IDs or gene symbols. Analyses were restricted to the Biological Process (BP) ontology. Genes expressed in iPSC-derived microglia or HMC3 cells served as the background set. Over-representation analysis was performed using a hypergeometric test, and p-values were adjusted using the Benjamini–Hochberg method (FDR < 0.05). Results were visualized with dot plots or bar plots.

### Transcription factor motif disruption and GRN inference

TF binding motifs were obtained from the HOCOMOCO^92^ human database. Variant–motif disruptions were evaluated using motifbreakR (v2.20.0)^93^. Only statistically significant motif hits were retained (filterp = TRUE), with a stringent p-value threshold of 1×10⁻⁴ to reduce false positives. The disruption score was calculated using the information content method, which accounts for the relative contribution of each nucleotide within the position weight matrix. To avoid bias from unequal nucleotide frequencies, a uniform background distribution of A, C, G, and T (0.25 each) was applied. Computations were parallelized using BiocParallel for efficiency.

For GRN construction, expression data from iMGL were filtered to include only genes expressed above 1 count per million (CPM) in at least 50% of samples, ensuring lowly expressed genes were excluded before normalization. Library sizes were adjusted using TMM normalization with edgeR^94^, and log-transformed expression values were obtained as log₂(CPM + prior count). GRNs were inferred using GENIE3 (v1.28.0)^95^, which applies a random forest–based approach with 1,000 decision trees to capture stable regulatory interactions. At each tree split, the number of candidate regulators was set to the square root of the total number of regulators, balancing bias and variance. A fixed random seed was used to ensure reproducibility. The top 500 regulator–target edges ranked by importance score were visualized in Cytoscape. Gene identifiers were harmonized across datasets using biomaRt (v2.60.0)^96,97^.

### Locus plot

The composite locus plot is done with the plot_grid function in the cowplot R package^98^. All plots were created using GRCh38 data, if data was originated from GRCh37, liftOver^99^ from rtracklayer (v1.64.0)^100^ was used for conversion before integration. GWAS summary statistics are from Bellengeuz et al^10^, and fine-mapping credible sets are from the SuSiE fine-mapping result. Microglia promoter and enhancer data are from the primary microglia ABC model built from Kosoy et al^14^. ATAC peak information is derived from ATAC sequencing data of iMGL. Gene models are from Ensembl (EnsDb.Hsapiens.v86, GRCh38)^101^ and labeled by HGNC gene symbol.

## DATA AVAILABILITY

Datasets used in this study are publicly available as follows: microglia eQTL summary statistics at Zenodo (https://zenodo.org/record/8250771) and microglia caQTLs at Synapse (https://doi.org/10.7303/syn26207321). Summary statistics from AD GWAS are available through the National Human Genome Research Institute-European Bioinformatics Institute GWAS catalog under accession numbers: GCST90027158, GCST90012877, and GCST90012878 (https://www.ebi.ac.uk/gwas/downloads/summary-statistics). List of variants tested for MPRA are available at: https://github.com/RajLabMSSM/AD_MPRA. All AD fine-mapped variants and emVars are also provided in Supplementary Tables.

## CODE AVAILABILITY

The code for the analyses described in this article can be found at: https://github.com/RajLabMSSM/AD_MPRA.

## ACKNOWLEDGMENTS

This work was supported in parts by grants from the National Institutes of Health (NIH)/National Institute on Aging (RF1AG065926, R01-AG065926, U01-AG068880, R21-AG091272, R01-NS116006, R21AG087875, R01AG050986, R01ES033630, TP220451). Any opinions, findings, and conclusions or recommendations expressed in this material are those of the authors and do not necessarily reflect the views of the NIH.

## AUTHORS’ CONTRIBUTIONS

CYL, TR and KJB conceived the study. AR and TR performed variant prioritization. CYL and KJB carried out the iMGL and HMC3 MPRA experiments. TZH conducted iMGL culture. CYL and LMH performed MPRA bioinformatics analyses. CYL, AR, LMH, TR and KJB wrote the original draft. All authors contributed to the review and editing of the manuscript.

