## Supplementary Figures for "Regulatory landscape of Alzheimer’s disease variants in human microglia"

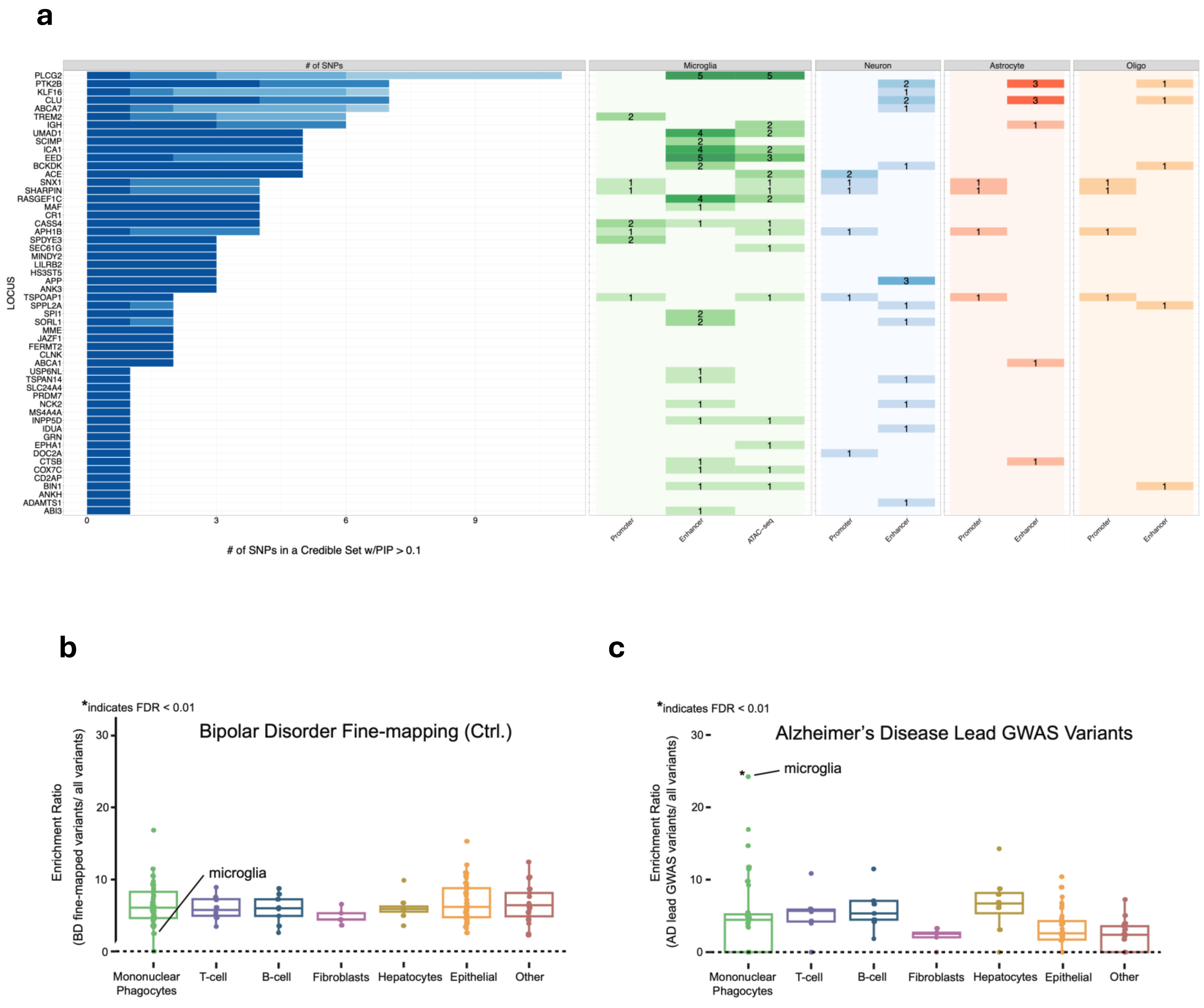


**Supplementary Figure 1. Summary of Fine-mapped Variants in Alzheimer's Disease (AD).**

(a) Fine-mapped results across all AD GWAS loci with the number of fine-mapped SNPs that overlap promoter and enhancer marks in microglia, neurons, astrocytes, and oligodendrocytes

(b) Enrichment ratio of bipolar disorder fine-mapped variants within enhancers identified by the Activity-by-Contact (ABC) model from 132 cell types as control

(c) Enrichment ratio of AD GWAS lead variants within enhancers identified by the Activity-by-Contact (ABC) model from 132 cell types

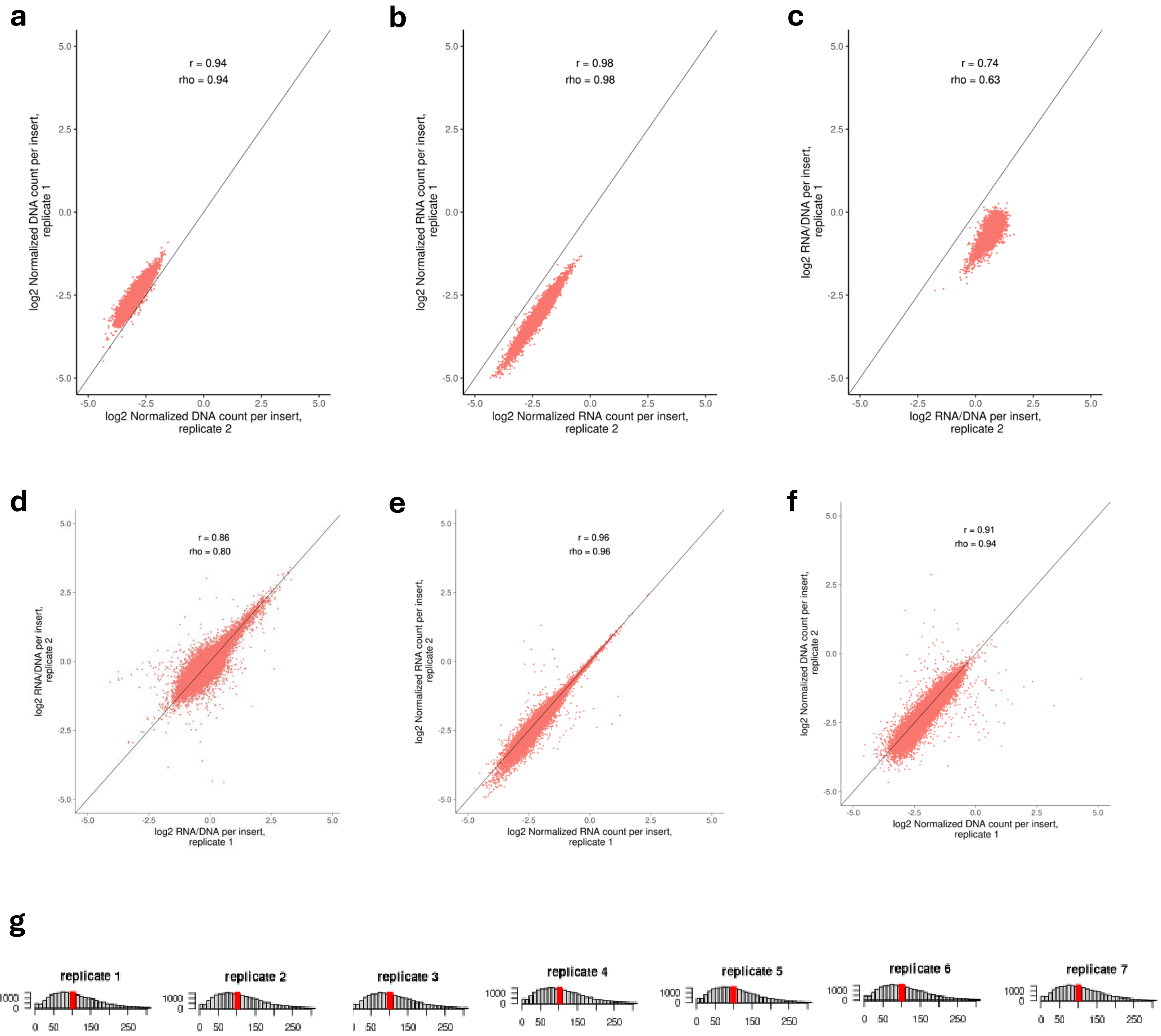


**Supplementary Figure 2. Summary of MPRA replicates correlation**

(a-c) DNA, RNA, log2 (RNA/DNA) of one pair of iMGL sample (replicate 1 and 2)

(d-f) DNA, RNA, log2 (RNA/DNA) of one pair of HMC3 sample (replicate 1 and 2)

(g) Detected barcode counts per replicate of iMGL sample

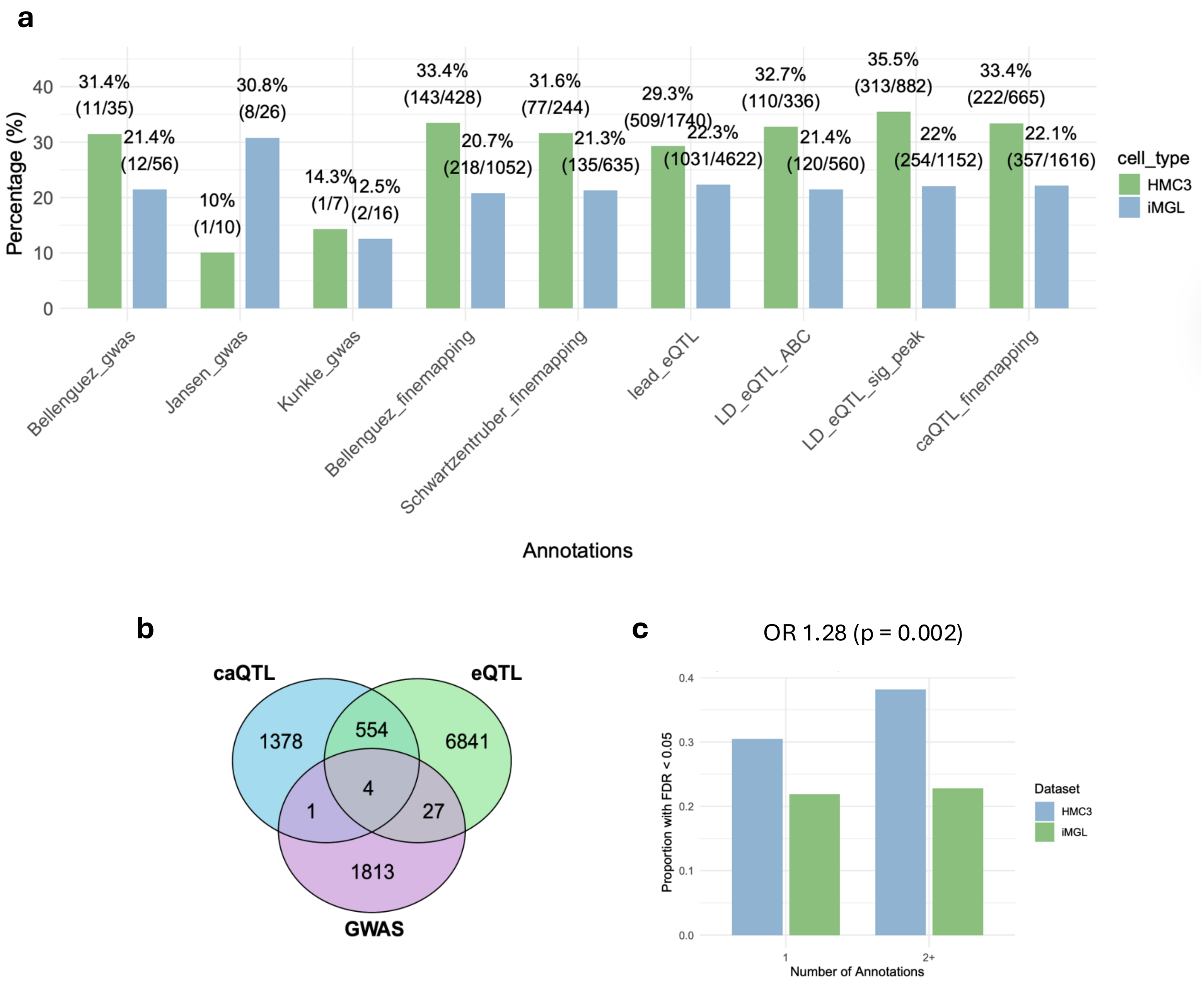


**Supplementary Figure 3. Functional annotation of regulatory variant predictions**

(a) Proportion of emVar classified by prioritization category. *_gwas indicates the lead SNPs identified in GWAS; *_finemapping indicates fine-mapped variants from GWAS loci; lead_eQTL denotes lead SNPs from the MiGA eQTL study; and LD_eQTL_* represents SNPs in high linkage disequilibrium with eQTL lead SNPs that also colocalize with ABC or ATAC annotations

(b) Overlap of variants successfully retrieved from iMGL samples among the three prioritization strategies

(c) Distribution of emVar based on the number of annotations each variant carries across GWAS, eQTL, and caQTL datasets

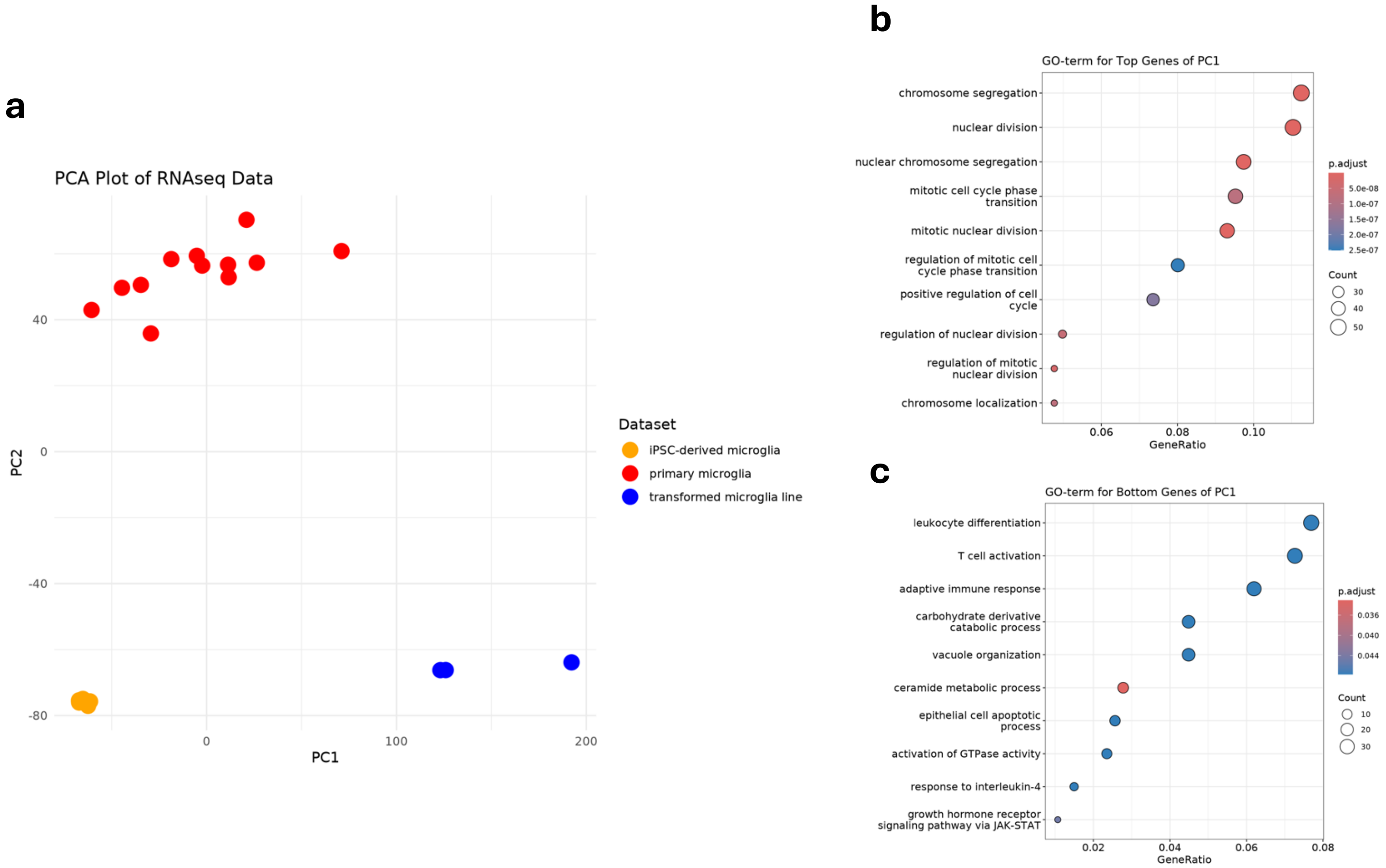


**Supplementary Figure 4. Transcriptional divergence among microglial models**

(a) Principal component analysis (PCA) of transcriptional profiles from primary microglia, iPSC-derived microglia, and the HMC3 microglial cell line (GSE219208).

(b) GO enrichment on the top 500 genes with the highest positive loadings

(c) the bottom 500 genes with the highest negative loadings in PC1

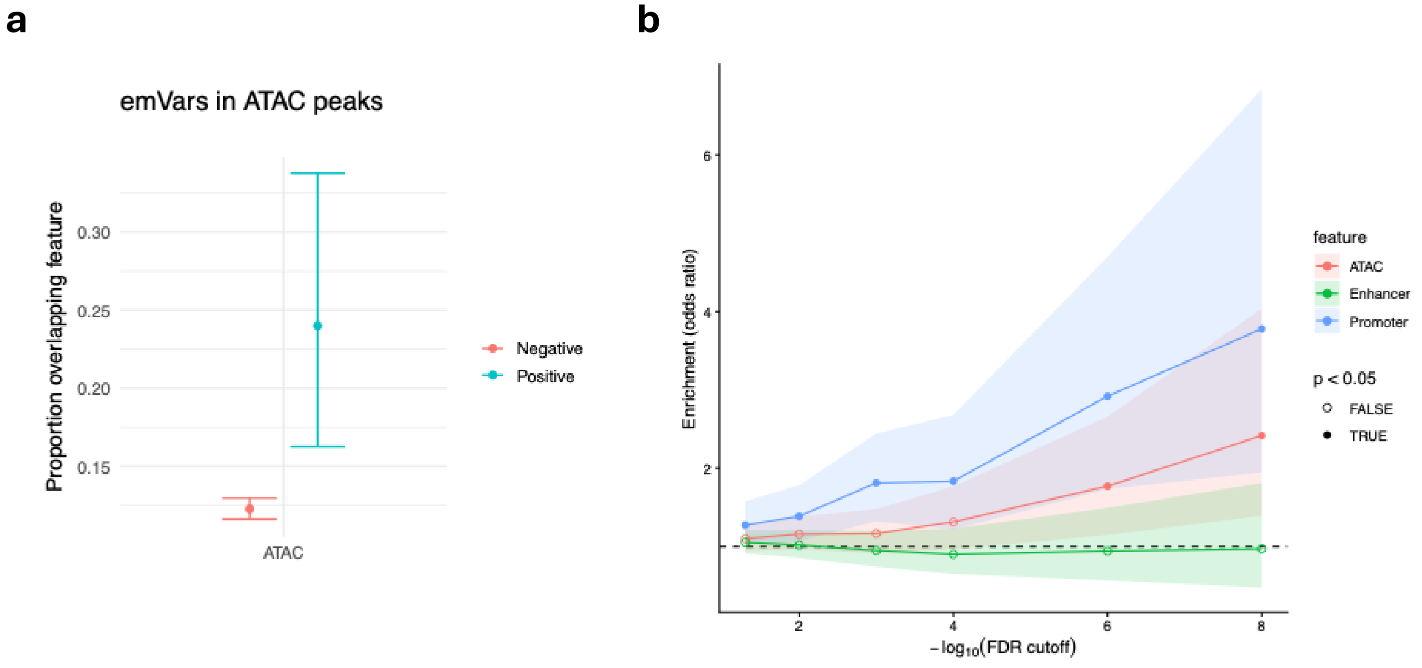


**Supplementary Figure 5. Enrichment of emVar within microglial regulatory annotations**

(a) Proportion of MPRA-positive ($p < 10^{-8}$) and MPRA-negative ($p > 10^{-8}$) variants overlapping iPSC-derived microglial ATAC-seq peaks

(b) Odds ratio of emVar enrichment within annotation evaluated under different MPRA significance cutoffs

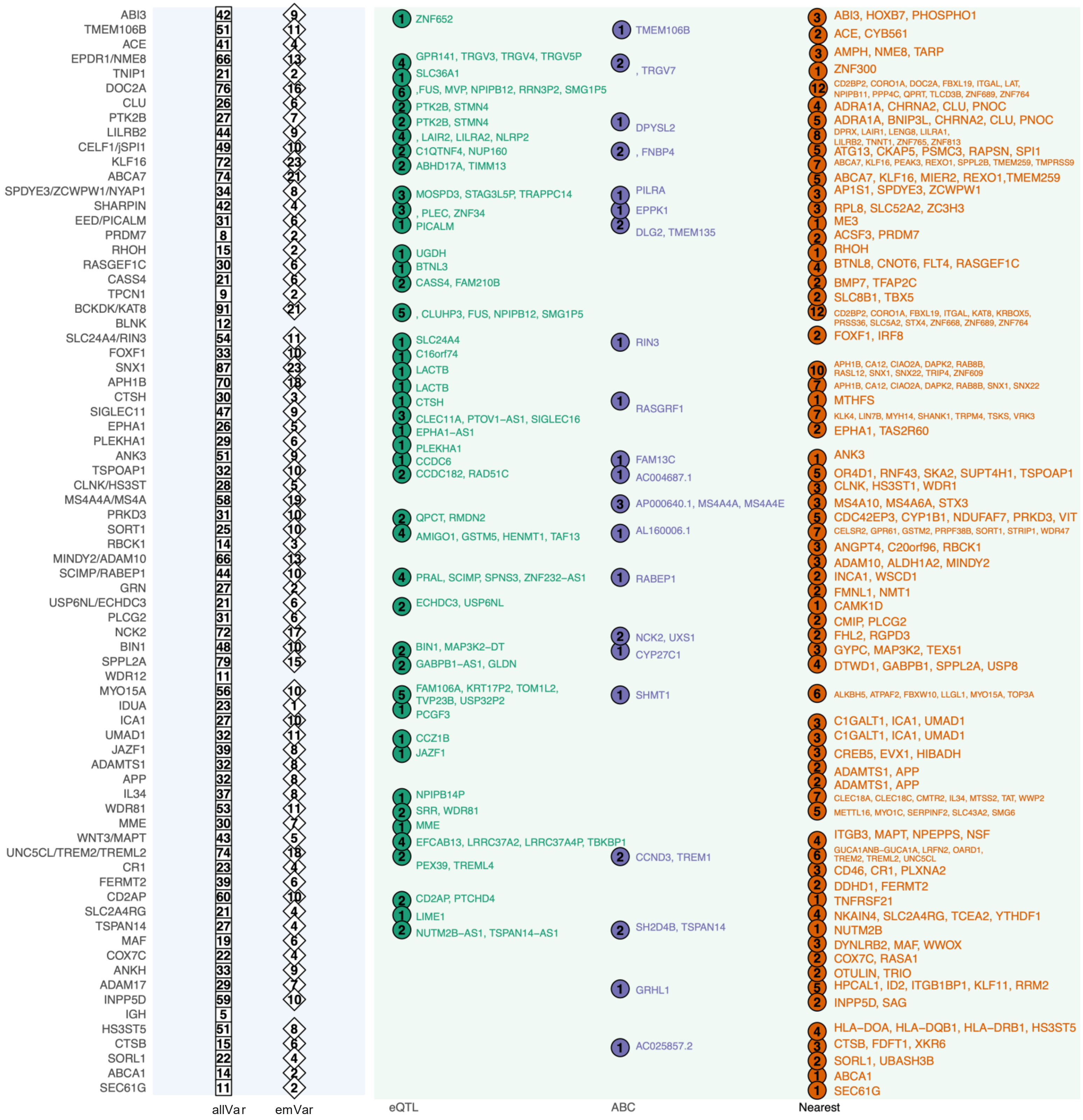

**Supplementary Figure 6. Summary of MPRA results across AD loci**

Overview of the number of all tested variants (allVar) and MPRA-identified functional regulatory variants (emVar), along with the genes linked to emVar through different variant-to-gene approaches (eQTL, ABC model, and nearest-gene)

**a**

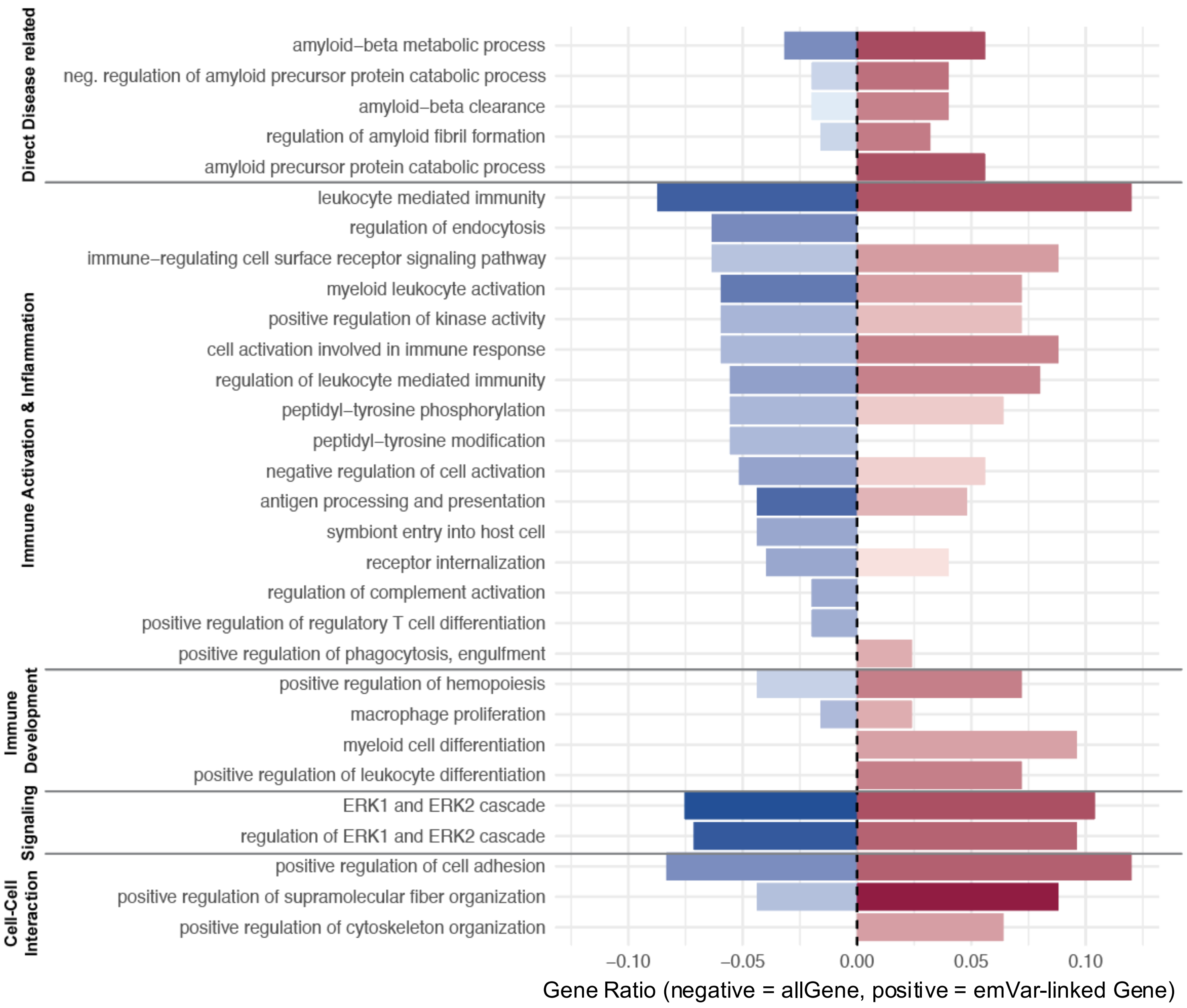


**Supplementary Figure 7. GO-term enrichment of allGene and emVar-mapped genes**

Comparison of GO-term enrichment for genes linked to all tested variants (allGene) versus mapped from emVar from AD GWAS prioritized variants

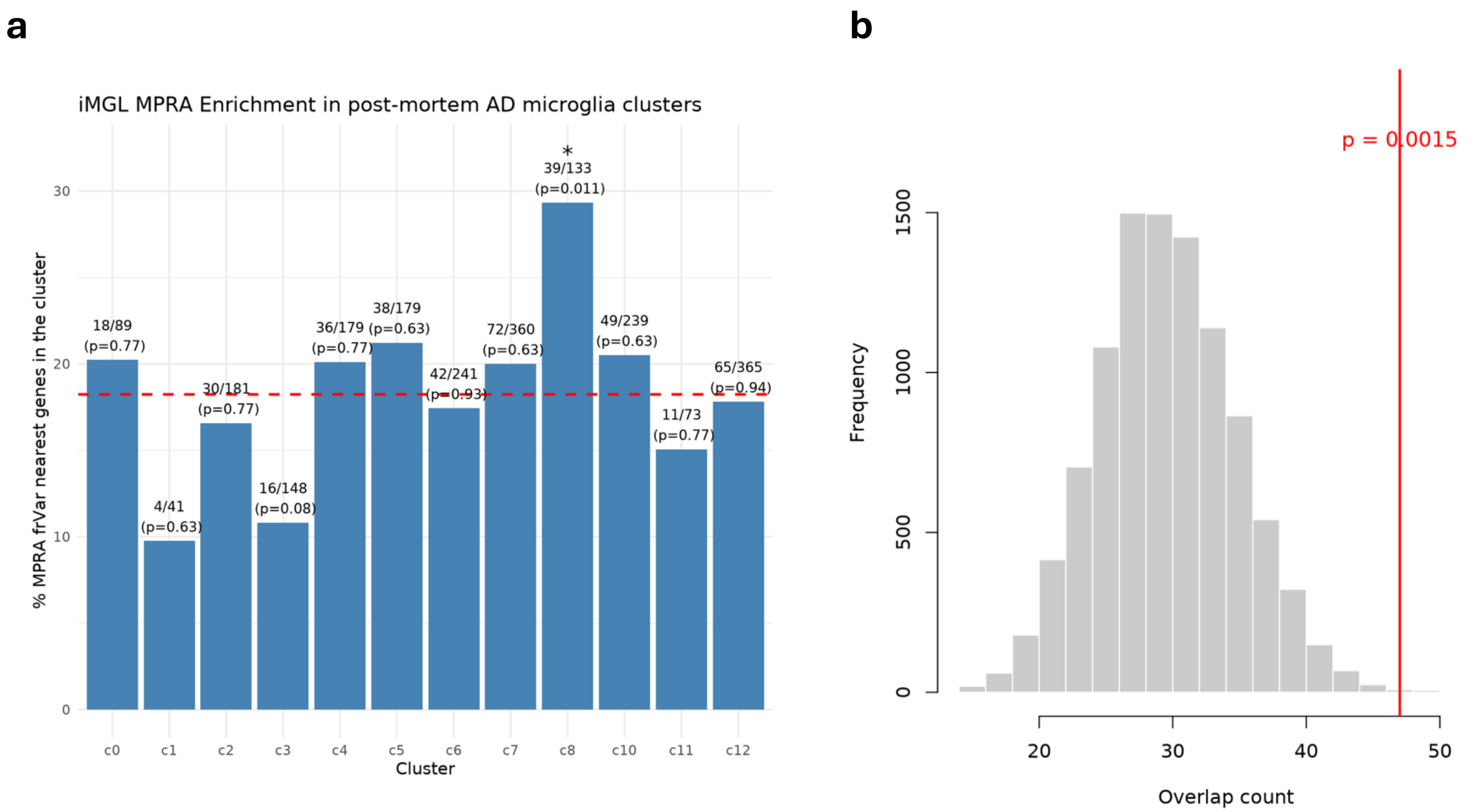


**Supplementary Figure 8. Overlap with AD differentially expressed genes (DEGs).**

(a) Overlap of emVar-linked genes with AD post-mortem microglial single-cell clusters

(b) Permutation on randomly selected genes as MPRA-linked genes (n=321) and overlap with AD post-mortem DEG.

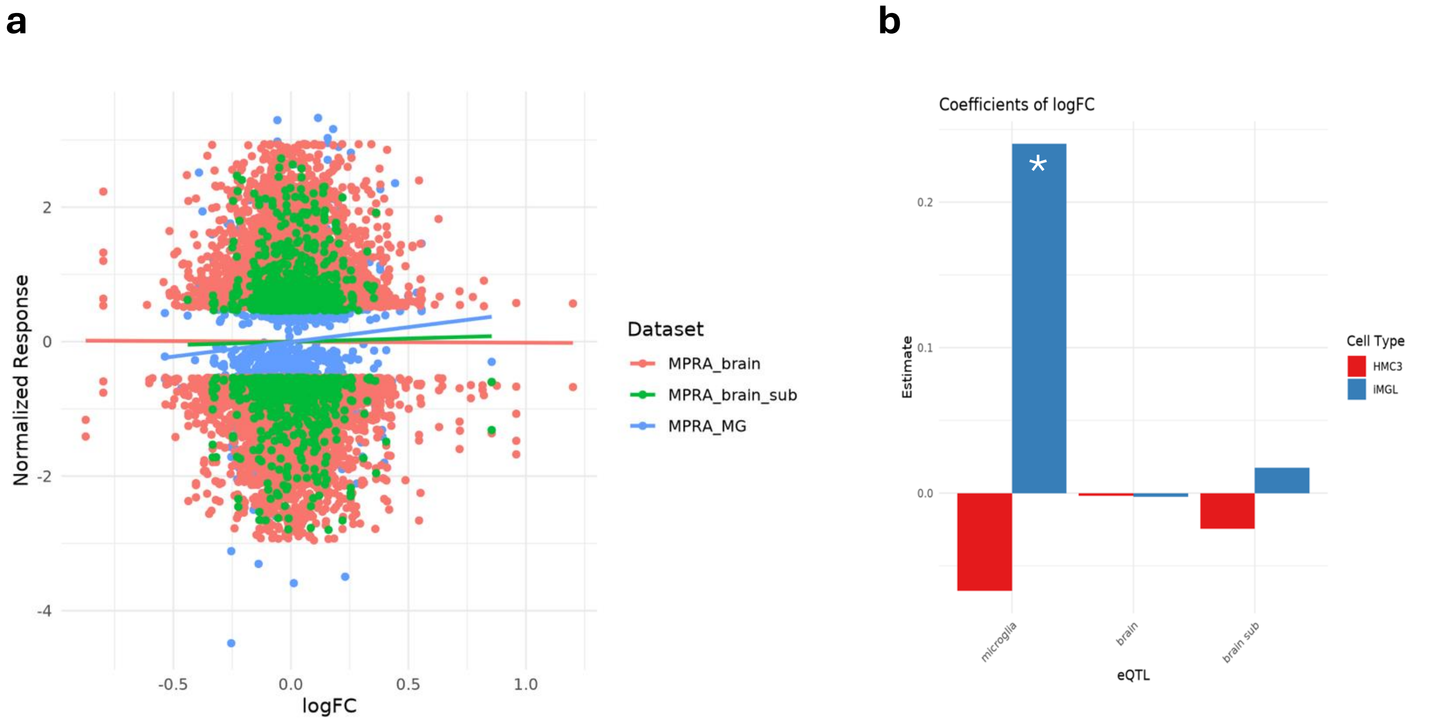


**Supplementary Figure 9. Correlation between MPRA effect sizes and eQTL effect estimates**

(a) Scatter plot of MPRA logFC versus normalized eQTL effect size across microglial (MPRA_MG), brain (MPRA_brain), and overlapping brain subset (MPRA_brain_sub) datasets

(b) Linear regression slope of correlation between MPRA logFC to microglial eQTL, brain eQTL, and subset of brain eQTL that overlapped with microglial eQTL

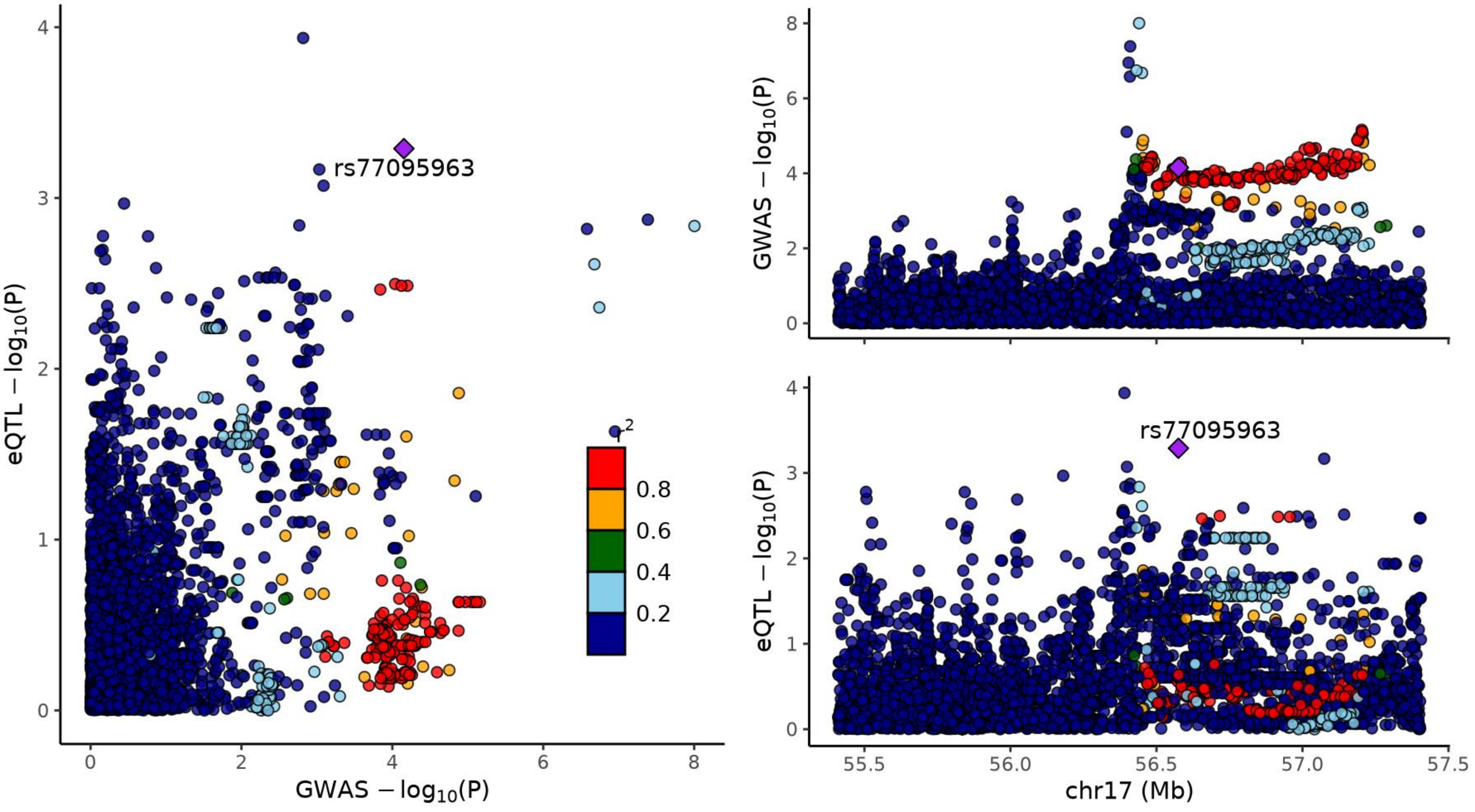


**Supplementary Figure 10. Colocalization between GWAS and eQTL signals at the TSPOAP1 locus**

Colocalization analysis between the Bellenguez et al. AD GWAS and microglial eQTLs revealed a Bayesian posterior probability of 55% for a shared causal signal (PP H4 = 0.55), 15% for distinct but significant signals (PP H3 = 0.15), and 29% for a GWAS-only signal (PP H1 = 0.29).
